# A Novel Strategy for Recurrent Heart Failure: Planned Hospitalization Before Clinical Worsening: A Retrospective Study of the Kurume-HEARTS Program

**DOI:** 10.64898/2026.04.01.26349992

**Authors:** Toshiyuki Yanai, Tatsuhiro Shibata, Kodai Shibao, Daiki Akagaki, Kota Okabe, Shoichiro Nohara, Jinya Takahashi, Koutatsu Shimozono, Yoshihiro Fukumoto

## Abstract

**Background:** The prevalence of heart failure (HF) is increasing worldwide, and rehospitalizations due to exacerbations remain a major clinical and economic burden. Beyond medical triggers, insufficient patient understanding and inadequate self-management often contribute to recurrent admissions. The Kurume-HEARTS program was developed to provide regular planned hospitalizations incorporating structured education, cardiac rehabilitation, and medication adjustment for patients with recurrent HF.

**Objective:** To retrospectively evaluate the clinical and economic impact of the Kurume-HEARTS program.

**Methods:** We enrolled consecutive patients with recurrent HF hospitalizations who underwent the program at Kurume University Hospital between January 2020 and October 2025. Outcomes compared planned versus unplanned hospitalizations within the same patients. Co-primary endpoints were total hospitalization cost and total length of stay per person-year. Secondary endpoints included per-hospitalization cost, length of stay, unplanned and planned admission frequency, and NT-proBNP levels at admission.

**Results:** Of 31 screened patients, 20 with recurrent heart failure were included. During a median follow-up of 27.1 months, 135 hospitalizations occurred (69 unplanned and 66 program-based). Total hospitalization cost per person-year was significantly lower during the Kurume-HEARTS program than during unplanned hospitalizations, while length of stay per person-year tended to be shorter. Per-admission cost and length of stay were significantly lower with the program, without differences in admission frequency. NT-proBNP levels at admission were higher during unplanned hospitalizations, indicating greater clinical instability.

**Conclusions:** The Kurume-HEARTS program can help reduce the cost and hospitalization length of unplanned admissions by enabling earlier intervention and structured inpatient management.

**Graphical Abstract:** **Concept of the Kurume-HEARTS program for preventing unplanned heart failure hospitalizations**

The upper panel illustrates conventional care, where patients with chronic heart failure frequently experience recurrent unplanned hospitalizations due to acute decompensation. The lower panel shows the planned strategy implemented in the Kurume-HEARTS program, in which patients undergo regularly planned hospitalizations for comprehensive multidisciplinary management before clinical worsening. This program includes patient education, nutritional guidance, medication optimization, exercise therapy, self-care instruction, and advance care planning. Through this proactive approach, the Kurume-HEARTS program may reduce the frequency/length of unplanned hospitalizations and hospitalization medical costs, contributing to more stable long-term disease management.

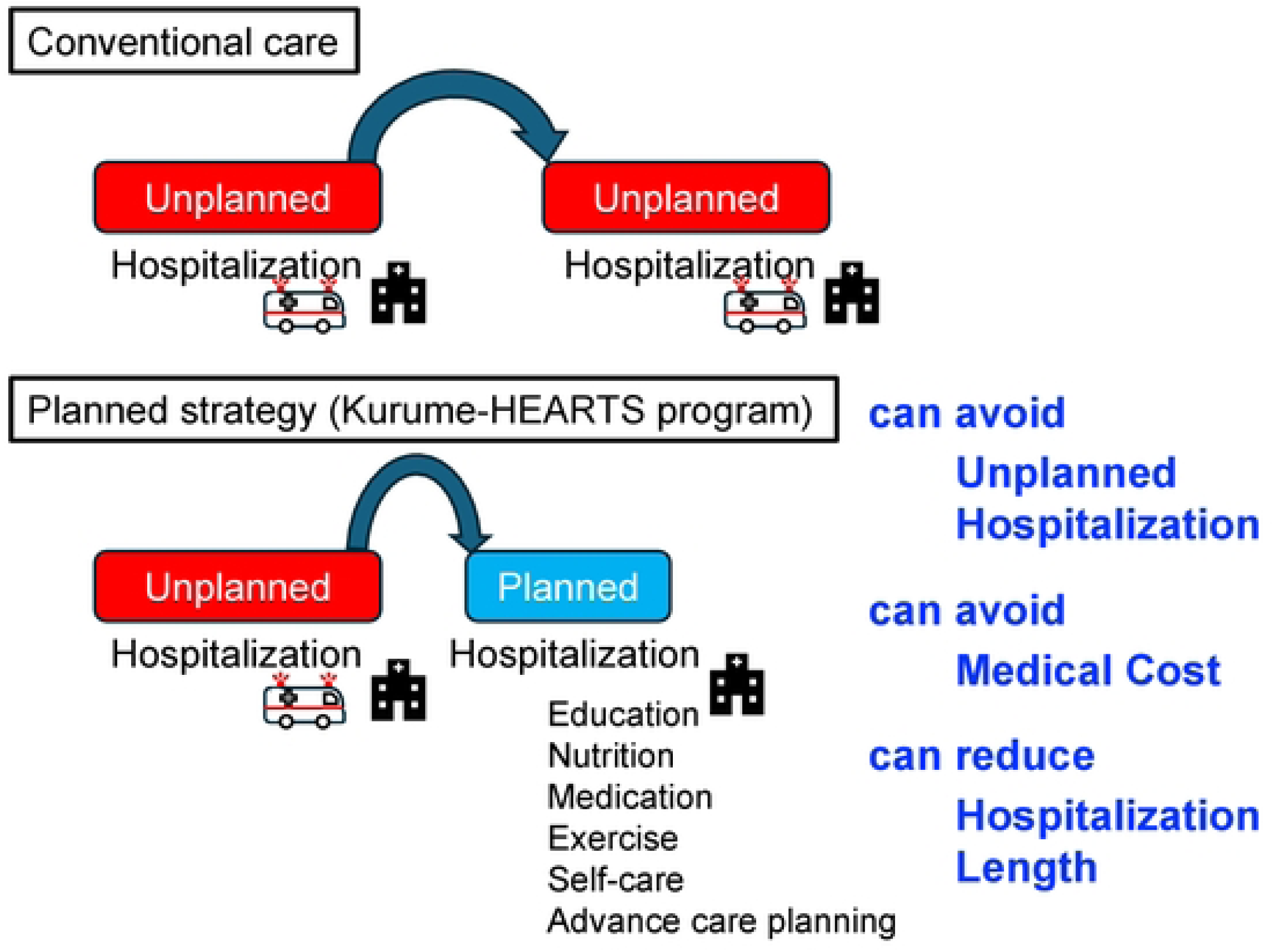

## Introduction

Heart failure (HF) affects more than 64 million individuals worldwide, and hospitalizations for decompensated HF contribute substantially to morbidity, mortality, and health care costs.[1, 2] In the United States alone, HF accounts for more than $30 billion in annual health care expenditures,[3] with inpatient care representing the largest component, primarily driven by comorbidities, invasive procedures, and readmissions.[4]

Despite advances in pharmacologic and non-pharmacologic therapies that improve survival in chronic HF,[5–9] acute decompensated heart failure (ADHF) remains one of the leading causes of hospitalization worldwide. Patients admitted with ADHF experience significant in-hospital morbidity and mortality, frequent rehospitalizations, and an elevated risk of subsequent cardiovascular death.[10] In the United States and Europe, there are more than 1 million hospitalizations for HF annually, with rehospitalization rates approaching one quarter of patients within 30 days.[11–14] Beyond their prognostic implications, rehospitalizations impose a disproportionate economic burden, with each recurrent admission adding substantially to cumulative costs. Rehospitalization for cardiovascular disease within 90 days of discharge is also independently associated with increased mortality risk, irrespective of the precise interval from discharge.[15] The prevalence of HF has risen sharply, particularly in Western countries, largely due to population aging.[16]

Effective inpatient care requires comprehensive diagnostic evaluation, triage and risk stratification, early initiation of guideline-directed medical therapy (GDMT), adequate decongestion, and structured discharge planning.[2, 17] In addition, patients with HF are encouraged to engage in daily self-care activities—collectively referred to as self-management—that help prevent clinical deterioration.[18] These strategies include adherence to prescribed pharmacotherapy, lifestyle modification, daily weight monitoring, and recognition and appropriate response to worsening symptoms.[18] Meta-analyses of interventions designed to promote self-management have consistently shown benefits, including reduced hospitalizations, improved quality of life (QoL), and decreased all-cause mortality.[19–21]

Self-management education should be tailored to the needs of specific populations, taking into account comorbid conditions as well as ethnic, social, cognitive, literacy, and cultural factors.[22] Hospitalization provides a valuable opportunity to deliver structured education; however, inpatient education models remain heterogeneous nationwide, with variable outcomes.[23] Meeting the educational needs of hospitalized patients will require sustained efforts, diverse strategies tailored to available resources, and further research to evaluate their impact on clinical and economic outcomes.[23] Nurse-led education programs, in particular, have been shown to improve QoL and significantly reduce HF-related hospitalizations, rehospitalizations, and mortality.[24, 25]

At our institution, we developed the Kurume Heart Education And Rehabilitation for Treatment Success (**Kurume-HEARTS**) program, a structured model of planned hospitalizations for HF education and rehabilitation targeting patients with chronic HF who experience recurrent hospitalizations due to acute exacerbations. This program specifically focuses on individuals who, despite receiving appropriate outpatient management—including optimized medical therapy, structured HF education, and cardiac rehabilitation—continue to experience repeated episodes of acute decompensation. Planned admissions are scheduled at the time of discharge, with the next hospitalization arranged in advance, thereby enabling regular reassessment, medication adjustment, and reinforcement of education. Based on our clinical experience, we hypothesize that the Kurume-HEARTS program—a proactive inpatient strategy—would be more effective than outpatient management in preventing acute exacerbations and reducing unplanned, resource-intensive hospitalizations. Therefore, the present study retrospectively evaluated whether the Kurume-HEARTS program reduces hospitalization costs and unplanned admission frequency in patients with recurrent HF. These patients represent a particularly challenging population in whom standard outpatient management alone has proven insufficient to prevent recurrent decompensation.

## Methods

### Study Design

This was a retrospective cohort study utilizing the database of the Department of Cardiovascular Medicine at Kurume University Hospital. We assessed the data in February 2026. The study population comprised patients who had experienced repeated hospitalizations for worsening HF. Eligible candidates included patients admitted with HF exacerbation who had a documented history of at least one HF-related hospitalization within the preceding year, counted from the index admission. We enrolled all consecutive patients who underwent planned hospitalizations for HF education and rehabilitation between January 2020 and October 2025. The present study received ethical approval from the Ethics Committee of Kurume University (approval no. 25235), and informed consent was waived because of the retrospective design; an opt-out approach was employed. The study was conducted in accordance with the ethical principles outlined in the Declaration of Helsinki.

### Study Population and Planned Hospitalization Program

Patients were eligible if they were aged ≥20 years, carried a diagnosis of HF, and met the following criteria: (1) at least one unplanned hospitalization for HF exacerbation within the previous 12 months, as defined by the Japanese Circulation Society (JCS 2017) /Japanese Heart Failure Society (JHFS 2017) guidelines;[26] (2) ability to perform activities of daily living independently; (3) ability to attend outpatient follow-up after discharge; (4) patients requiring enhanced self-care support (e.g., poor medication adherence, inadequate weight management, difficulty adhering to dietary recommendations, or insufficient social support such as living alone).

The interval between planned hospitalizations was set shorter than the patients’ prior recurrence interval of HF exacerbation. For example, patients typically re-hospitalized every 6 months were scheduled for planned hospitalizations every 4 months (**Figure 1A top**). Patients deemed to require re-education were scheduled for Kurume HEARTS program at 6- or 12-month intervals, with timing tailored to individual clinical status (**Figure 1A bottom**).

**Figure 1.**
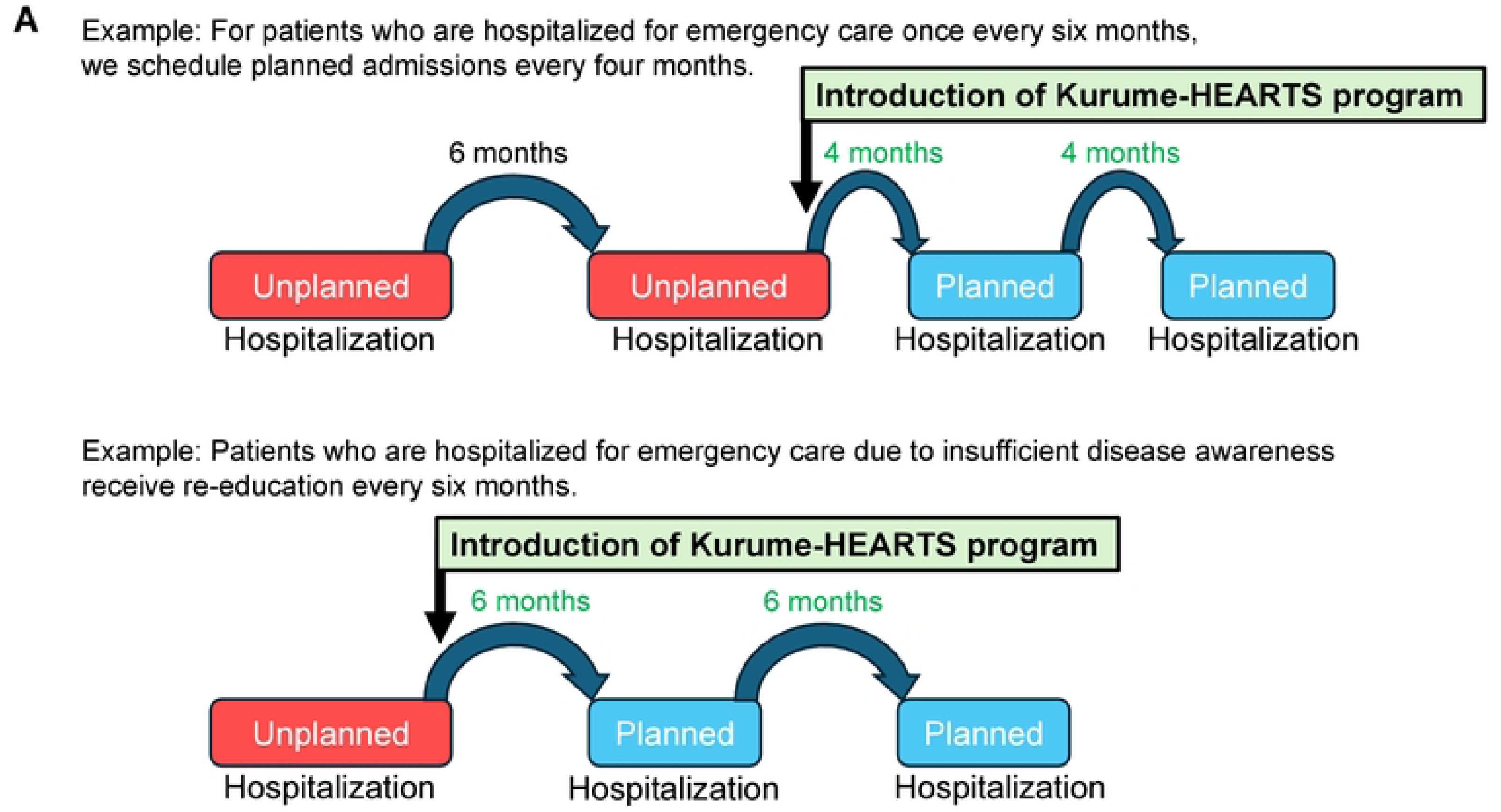
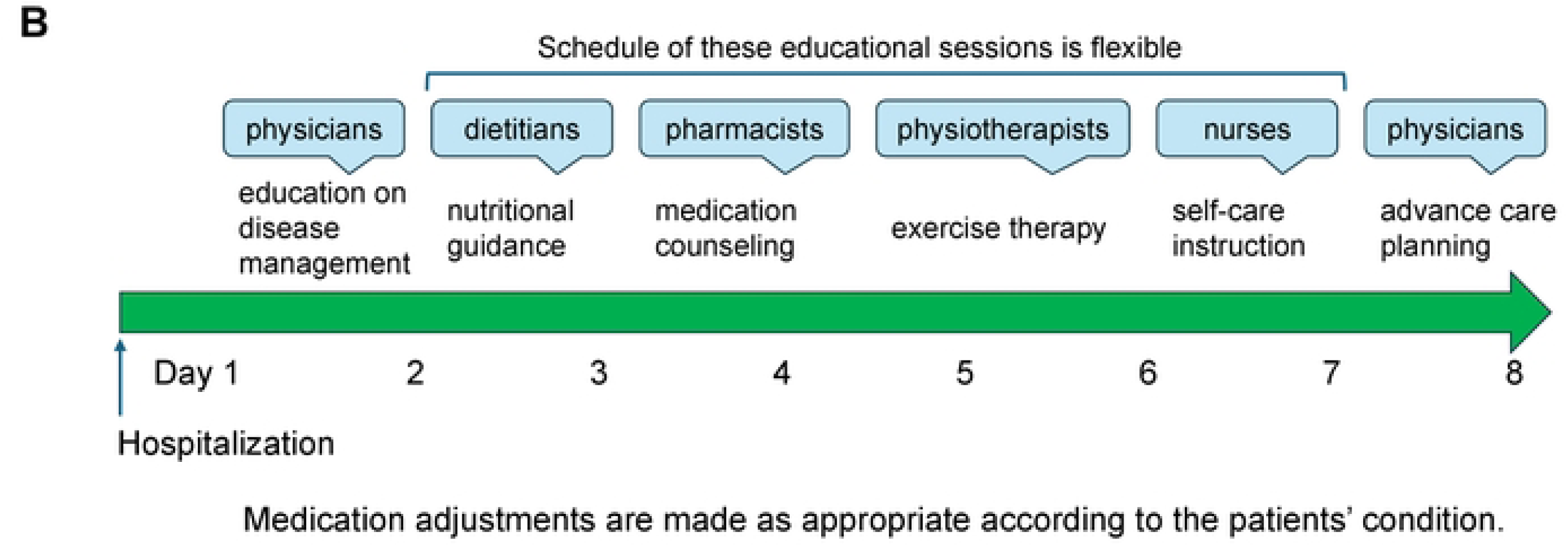
Examples of Kurume-HEARTS program at our institution. (A) Implementation scheme for planned hospitalizations (Kurume-HEARTS program) for heart failure (HF) education and rehabilitation. For patients who typically require unplanned hospitalization every 6 months (top), planned hospitalizations are arranged every 4 months to prevent acute deterioration. For patients admitted emergently due to limited disease awareness (bottom), re-education is provided every 6 months. (B) Overview of the Kurume-HEARTS program for HF education and rehabilitation. During an approximately 8-day hospitalization, multidisciplinary education is delivered with flexible scheduling of sessions: physicians (disease management and advance care planning), dietitians (nutritional counseling), pharmacists (medication guidance), physiotherapists (exercise therapy), and nurses (self-care training). Medication adjustments are tailored to each patient’s clinical status.

Our Kurume-HEARTS program comprised a multidisciplinary approach: physicians provided education on disease management, dietitians offered nutritional counseling, pharmacists delivered medication guidance, physiotherapists conducted exercise therapy, nurses provided instruction in self-care, and physicians additionally addressed advance care planning. Educational sessions were scheduled flexibly, and medications were adjusted as clinically appropriate during the hospitalization (**Figure 1B**). Exclusion criteria were: (1) diagnosis of dementia; (2) clinical determination by the attending physician that participation was inappropriate; (3) scheduled surgery; (4) initiation of or immediate post-initiation period of dialysis; (5) terminal stage of malignant diseases.

Among consecutive eligible patients, hospitalization costs before and after initiation of Kurume-HEARTS program were analyzed during the study period.

### Data Collection

Data were extracted from the institutional database and included baseline demographic and clinical characteristics: age, sex, height, weight, systolic and diastolic blood pressure, pulse rate, medication use, and planned or unplanned hospitalization. Body mass index (BMI) was calculated as weight in kilograms divided by height in meters squared.

Laboratory data were obtained from peripheral venous blood samples analyzed at the Kurume University Hospital central laboratory, including white blood cell count, hemoglobin, hemoglobin A1c, serum creatinine, serum sodium, and N-terminal pro-B-type natriuretic peptide (NT-proBNP). Additional data included the presence or absence of congestion on chest radiography and left ventricular ejection fraction (LVEF), measured using the two-dimensional modified Simpson’s method.

HF was classified according to the underlying etiology, including ischemic heart disease, cardiomyopathy, valvular heart disease, and pulmonary arterial hypertension. Cardiomyopathies included amyloidosis, hypertrophic cardiomyopathy, and dilated cardiomyopathy. Valvular disease was diagnosed in patients with clinically significant lesions of the aortic, mitral, tricuspid, or pulmonary valves (e.g., aortic stenosis, aortic regurgitation, mitral regurgitation).

### Calculation of Hospitalization Costs in Japan Using the Diagnosis Procedure Combination System

In Japan, hospitalization costs at acute care hospitals adopting the Diagnosis Procedure Combination (DPC) system are determined through a mixed-case payment framework that integrates per-diem bundled payments with fee-for-service components. The DPC system, implemented nationwide since 2003, was introduced to standardize reimbursement, enhance transparency, and encourage efficiency in inpatient care.

In this study, hospitalization costs were calculated according to the DPC system, encompassing hospitalization charges, medical management fees, injection fees, diagnostic tests, surgical procedures, imaging, rehabilitation, comprehensive medical treatment fees, advanced diagnostic examinations, and high-cost interventions (e.g., acute-phase treatment of ADHF, cardiac catheterization, device implantation). Importantly, oral medication fees during hospitalization and discharge prescription fees were excluded from the calculation in our analyses.

### Endpoints

This study compared outcomes between unplanned hospitalizations and admissions for the Kurume-HEARTS program using a paired, within-patient design. The co-primary endpoints were total hospitalization cost per patient and total hospitalization length of stay per patient during the study period. Secondary endpoints included per-hospitalization cost per patient, per-hospitalization length of stay per patient, number of admissions per person-year, and NT-proBNP per admission per patient during the study period.

### Statistical Analysis

Continuous variables are presented as mean ± standard deviation (SD) or median and interquartile range (IQR). The primary analysis was a paired, within-patient comparison of outcomes between unplanned hospitalizations and admissions for the Kurume-HEARTS program. All paired comparisons were performed using the Wilcoxon signed-rank test. For the primary endpoints, the observation period for each patient was calculated from their first admission date to their last discharge date. The total cost and total length of stay for each hospitalization type were then annualized by dividing by this observation period (in years) to yield cost per person-year and length of stay per person-year, respectively. The effect size for these primary endpoints is reported as the Hodges-Lehmann median of the within-patient differences (Unplanned hospitalizations − Kurume-HEARTS program) with its 95% confidence interval (CI).

For the secondary endpoints, per-hospitalization cost was summarized as the patient-level geometric mean for each hospitalization type, while per-hospitalization length of stay was computed as the patient-level median. The effect sizes for both endpoints are reported as the Hodges-Lehmann median of the within-patient differences. NT-proBNP was summarized as the patient-level geometric mean; however, the comparison was performed on the log-transformed values to derive the geometric mean ratio (GMR = Unplanned hospitalizations / Kurume-HEARTS program) and its 95% CI. The number of admissions was annualized to derive admissions per person-year, and the effect size is reported as the median of the within-patient differences.

A two-sided p-value of < 0.05 was considered statistically significant for all analyses. All statistical analyses were performed using R version 4.5.1 (R Foundation for Statistical Computing, Vienna, Austria).

## Results

### Patient Characteristics Enrolled in Kurume-HEARTS program

During the study period, 31 patients were initially screened and enrolled. Of these, 6 patients who did not experience unplanned hospitalization for HF during the study period, 4 patients who were unable to initiate planned hospitalization due to frequent HF exacerbations, and 1 patient who declined outpatient follow-up were excluded (**Figure 2**). Consequently, a total of 20 patients (13 men, 7 women) with a mean age of 69.2 years were included in the final analysis (**Table 1**). Underlying conditions were heterogeneous, including ischemic (n=4), non-ischemic cardiomyopathy (n=10; 4 dilated, 3 hypertrophic cardiomyopathy, 3 cardiac amyloidosis), valvular heart disease after surgery (n=4), and right heart failure due to pulmonary arterial hypertension (n=2). At first planned admission, mean left ventricular ejection fraction was reduced (39.9%), mean NT-proBNP was elevated (5363 pg/mL), and renal function was impaired (mean eGFR as low as 39.4 mL/min/1.73 m²) (**Table 1**). Anemia and dyslipidemia were also common (**Table 1**).

**Figure 2.**
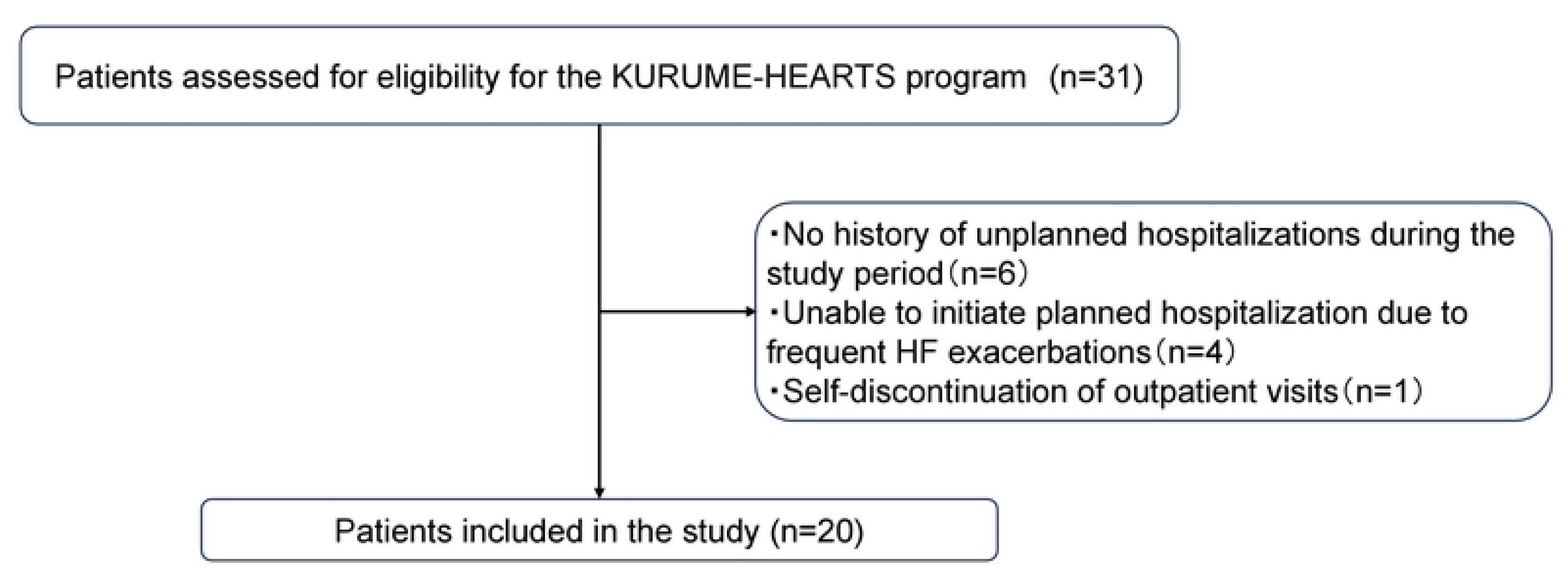
Study enrollment flowchart. Of 31 patients assessed for eligibility for the KURUME-HEARTS program, 11 were excluded (no history of unplanned hospitalization during the study period, n=6; inability to initiate planned hospitalization due to frequent heart failure exacerbations, n=4; self-discontinuation of outpatient visits, n=1). The final study cohort comprised 20 patients.

**Table 1.** Patients characteristics.

| **At First Planned Admission** |  | (n=20) |
| --- | --- | --- |
| Sex |  |  |
|  | Male (n, %) | 13, 65% |
|  | Female (n, %) | 7, 35% |
| Underlying heart disease |  |  |
|  | Ischemia (n, %) | 4, 20% |
|  | Non-ischemic cardiomyopathy (n, %) | 10, 50% |
|  | Valvular heart disease (n, %) | 4, 20% |
|  | Pulmonary arterial hypertension (n, %) | 2, 10% |
| Age at first planned admission (years) |  | 69.2 ± 15.3 |
| Systolic blood pressure (mmHg) |  | 99.6 ± 21.3 |
| Diastolic blood pressure (mmHg) |  | 62.0 ± 13.8 |
| Heart rate (bpm) |  | 73.0 ± 9.6 |
| Body mass index |  | 23.0 ± 5.2 |
| Left ventricular ejection fraction (%) |  | 39.9 ± 19.2 |
| NT-proBNP (pg/ml) |  | 5363 ± 5569 |
| eGFR (ml/min/1.73 m<sup>2</sup>) |  | 39.4 ± 20.7 |
| Hemoglobin (g/dl) |  | 14.3 ± 9.7 |
| LDL-C (mg/dl) |  | 85.8 ± 39.2 |
| **In-hospital death** |  |  |
|  | At planned hospitalization (n,%) | 2, 10% |
|  | At unplanned hospitalization(n, %) | 3, 15% |
| **At Discharge from Final Planned Admission** |  | (n=18) |
|  | RAAS inhibitors (n, %) | 15, 83% |
|  | β-blockers (n, %) | 16, 89% |
|  | MRAs (n, %) | 15, 83% |
|  | SGLT2 inhibitors (n, %) | 17, 94% |
|  | Diuretics (n, %) | 16, 89% |
|  | Device therapy (n, %) | 13, 72% |

Continuous variables are expressed as the mean ± standard deviation, categorical variables as n, %. eGFR, estimated glomerular filtration rate; NT-proBNP, N-terminal pro-B-type natriuretic peptide; LDL-C; low-density lipoprotein cholesterol; MRA, mineralocorticoid receptor antagonist; RAAS, renin-angiotensin-aldosterone system; SGLT2, sodium-glucose cotransporter 2.

During the first planned hospitalizations of Kurume-HEARTS program, hemodynamic parameters stabilized. At final discharge after the program, medical therapy was optimized: renin-angiotensin-aldosterone system (RAAS) inhibitors, including sacubitril/valsartan, in 15 patients, β-blockers in 16, mineralocorticoid receptor antagonists (MRAs) in 15, and sodium-glucose cotransporter 2 (SGLT2) inhibitors in 17; diuretics in 16, and 13 had device therapy (pacemaker or cardiac resynchronization therapy defibrillator) (**Table 1**).

### Primary Outcomes

During a median follow-up period of 27.1 months [13.7-45.5], a total of 135 hospitalizations from 20 patients were included in the analysis. Of these admissions, 69 were classified as unplanned and 66 were for the Kurume-HEARTS program. The median total hospitalization cost per patient was 2,589 [IQR 1,416–4,766] (×1,000 JPY) for unplanned hospitalizations versus 1,014 [IQR 697–1,857] (×1,000 JPY) for Kurume-HEARTS program (median difference +1,658 (×1,000 JPY); 95% CI: 178 to 3,487; p=0.015) during the study period **(Table 2**, **Figure 3)**. The median total hospitalization length of stay per patient was 47.4 [IQR 22.6–63.2] days for unplanned versus 19.9 [IQR 12.5–41.5] days for Kurume-HEARTS program (median difference +19.3 days; 95% CI: -1.1 to 41.9; p=0.070) **(Table 2**, **Figure 3)**.

**Figure 3.**
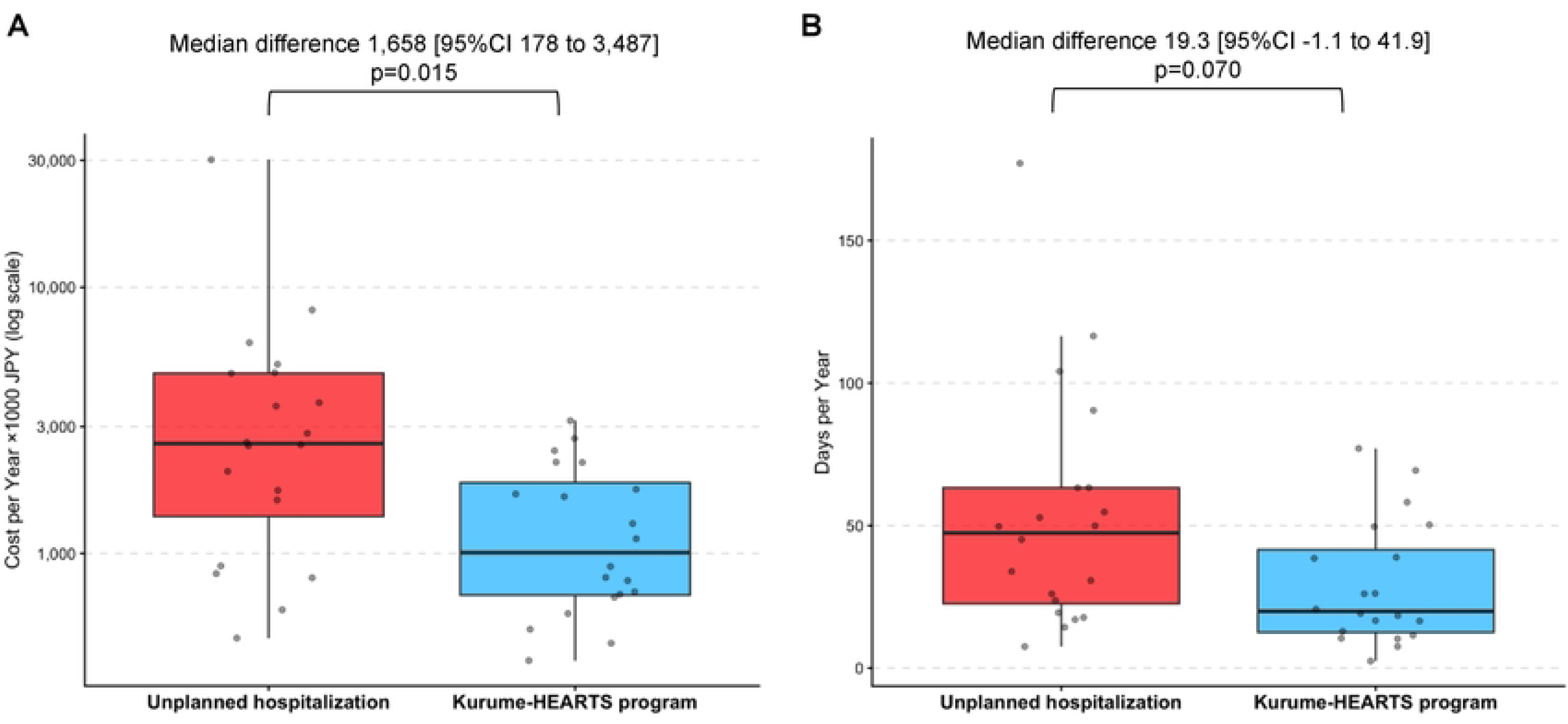
Total hospitalization cost and length of stay per person-year. Comparison of healthcare utilization between unplanned hospitalizations and the KURUME-HEARTS program. (A) Total hospitalization cost per person-year (log scale). The median annual hospitalization cost was significantly lower during participation in the KURUME-HEARTS program compared with unplanned hospitalization. (B) Total length of hospital stay per person-year. The KURUME-HEARTS program was associated with a numerically shorter annual hospitalization duration compared with unplanned hospitalization.

**Table 2.**
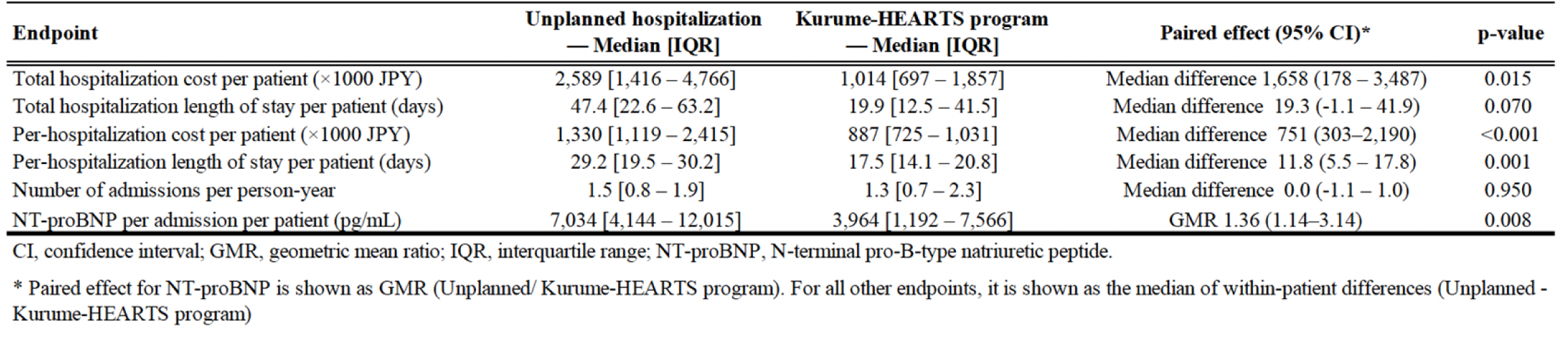
Paired Analysis of Outcomes for Unplanned Hospitalization versus Kurume-HEARTS Program (N=20)

### Secondary Outcomes

In the paired, within-patient analysis, the per-hospitalization cost per patient was significantly higher for unplanned hospitalizations. Median difference was 751 (95% CI, 303–2,190; p<0.001), with per-hospitalization cost per patient of 1,330 [IQR, 1,119–2,415] (×1,000 JPY) for unplanned and 887 [IQR, 725–1,031] (×1,000 JPY) for Kurume-HEARTS program admissions **(Table 2**, **Figure 4)**. The per-hospitalization length of stay per patient was also significantly longer for unplanned hospitalizations. The median within-patient difference was +11.8 days (95% CI, 5.5–17.8; p=0.001); patient-level medians were 29.2 [IQR, 19.5–30.2] days for unplanned versus 17.5 [IQR, 14.1–20.8] days for Kurume-HEARTS program admissions **(Table 2**, **Figure 4)**. There was no significant difference in the number of admissions per person-year (median difference, 0.0; 95% CI, -1.1–1.0; p=0.950) **(Table 2)**. NT-proBNP per admission per patient was significantly higher during unplanned hospitalizations compared to Kurume-HEARTS program admissions (GMR, 1.36; 95% CI, 1.14–3.14; p=0.008) **(Table 2)**.

**Figure 4.**
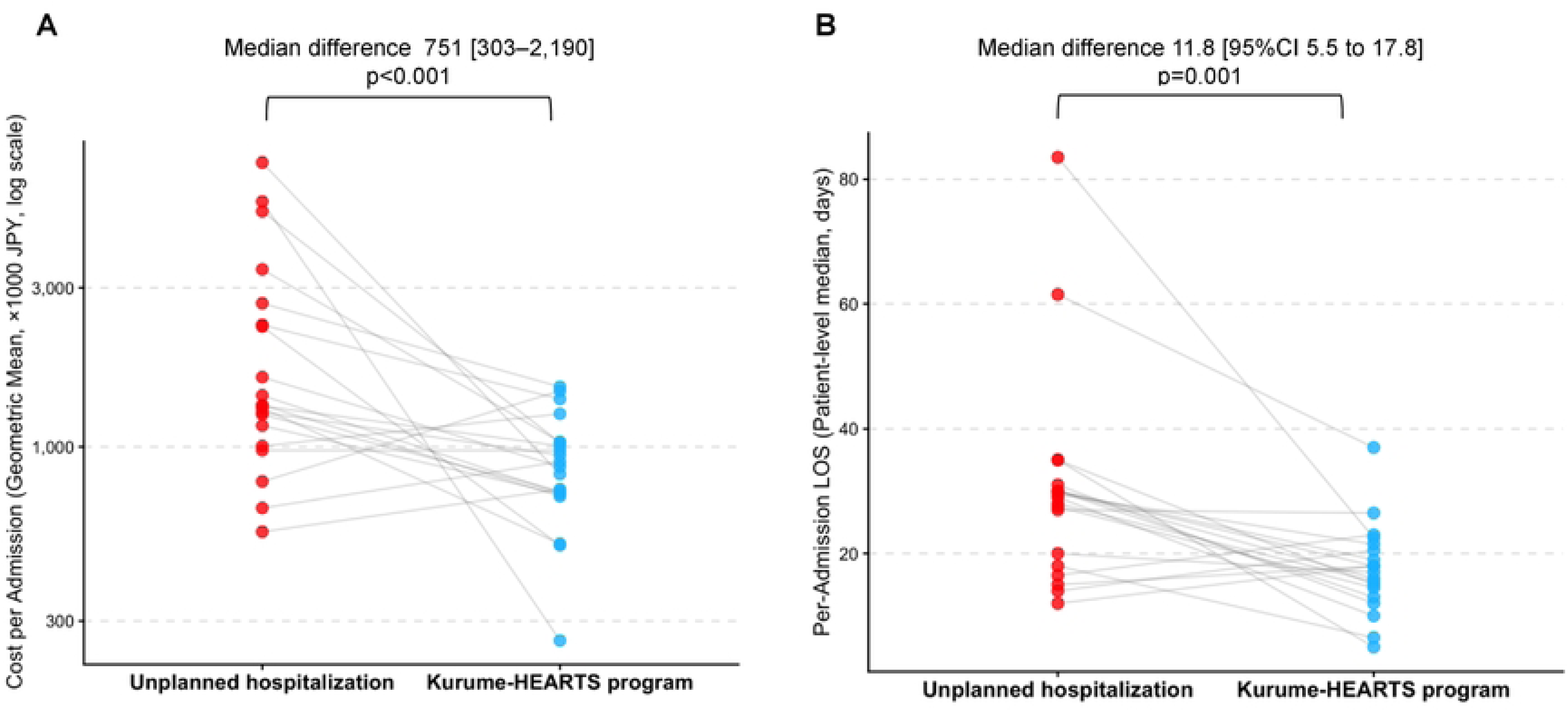
Per-hospitalization cost and length of stay per patient. Patient-level comparison of hospitalization characteristics between unplanned hospitalization and the KURUME-HEARTS program. (A) Cost per admission (log scale, geometric mean ×1000 JPY). The KURUME-HEARTS program was associated with significantly lower per-admission cost compared with unplanned hospitalization. (B) Per-admission length of stay (patient-level median days). Hospitalizations during the KURUME-HEARTS program were significantly shorter than unplanned hospitalizations.

## Discussion

The present study evaluated the clinical and economic impact of the Kurume-HEARTS program, a structured planned hospitalization strategy for patients with recurrent HF, using a within-patient paired analysis comparing unplanned hospitalizations with program-based admissions. The principal findings of this study are as follows. First, the Kurume-HEARTS program was associated with significantly lower per-hospitalization cost and shorter length of stay compared with unplanned HF hospitalizations. Second, although the total cost per person-year was numerically lower and the total length of stay per person-year tended to be shorter during participation in the program, the overall hospitalization burden per person-year did not significantly differ between strategies. Third, unplanned hospitalizations occurred under conditions of greater clinical instability, as reflected by significantly higher NT-proBNP levels at admission. Finally, following participation in the Kurume-HEARTS program, optimization of GDMT was confirmed, including the use of RAAS inhibitors, β-blockers, MRAs, and SGLT2 inhibitors, as well as device therapy when indicated. Collectively, these findings suggest that a structured planned hospitalization program for patients with advanced HF may improve the efficiency of inpatient care and reduce the clinical and economic burden of individual HF admissions.

### Burden of Unplanned Hospitalizations

Unplanned hospitalizations are a principal driver of resource use and costs in HF, which is the leading cause of hospitalization among older adults in the United States.[27] Early post-discharge is a particularly vulnerable period; contemporary cohorts report ≈30% risk of rehospitalization or all-cause death within 6 months of an HF admission.[28, 29] Economic evaluations (e.g., REHAB-HF) highlight that most direct costs (>75%) accrue to inpatient care, and that even when interventions improve function and QoL, inpatient expenditures remain substantial.[30–32]. Our findings mirror this clinical reality: patients admitted emergently typically present with advanced congestion and organ dysfunction, necessitating intensive treatment, prolonged monitoring, and complex management, all of which lengthen stays and inflate costs.

### Value of Planned Hospitalizations of Kurume-HEARTS program

This study demonstrated that a planned, structured hospitalization strategy may provide an additional framework for optimizing care in patients with recurrent HF. Multidisciplinary HF disease management and cardiac rehabilitation programs have consistently been shown to reduce HF readmissions and improve functional capacity and QoL through comprehensive interventions including medication optimization, patient education, exercise training, and coordinated follow-up. However, many patients with advanced or unstable HF continue to experience repeated unplanned hospitalizations despite these strategies. The Kurume-HEARTS program represents a complementary approach in which hospitalization itself is used proactively as a platform for clinical stabilization, reassessment of hemodynamics and volume status, and systematic optimization of GDMT.

By scheduling hospitalizations proactively, multidisciplinary teams can titrate therapies, reinforce self-care behaviors, and address emerging clinical issues before severe decompensation occurs. In our cohort, planned hospitalizations were consistently associated with lower per-admission costs and shorter lengths of stay compared with unplanned HF admissions. Although these scheduled admissions may modestly increase the number of hospital encounters, their lower intensity and shorter duration appeared to offset the clinical and economic burden associated with emergency hospitalizations.

In practice, patients enrolled in the Kurume-HEARTS program typically undergo repeated planned hospitalizations over time for HF education, rehabilitation, and medication optimization. The optimal duration and frequency of such planned admissions remain uncertain and may require individualized adjustment according to patient characteristics. Currently, the program duration is typically set at 5–10 days, but defining the most effective schedule represents an important area for future investigation. Furthermore, as the program becomes more widely implemented, the accumulation of multidisciplinary educational experience among healthcare staff may further enhance the quality of patient education and disease management. Such system-level learning effects may ultimately extend the benefits of the program beyond enrolled patients, suggesting a broader and potentially sustainable impact on HF care delivery.

### Integration Into Heart Failure Programs

Planned hospitalizations for HF education and rehabilitation should be considered within broader multidisciplinary programs that combine education, dietary and exercise counseling, medication titration, and transitions-of-care support. Evidence from telerehabilitation (e.g., TELEREH-HF) suggests improvements in functional status and QoL, though durable reductions in hard outcomes may require sustained, structured follow-up beyond short-term programs.[33–36]. At the same time, the TELEREH-HF study demonstrates that the improvements achieved during the 9-week hybrid comprehensive telerehabilitation do not increase the percentage of days alive and out of the hospital or reduce mortality or hospitalization rates on longer-term follow-up (after the intervention was stopped).[33] Education-centered, multidisciplinary strategies—self-management training on medication adherence, volume control, diet, weight monitoring, and early symptom response—improve knowledge, confidence, and clinical endpoints, and reduce unplanned visits and hospitalizations. Planned hospitalizations offer a practical platform to deliver these interventions intensively and consistently.

### Policy and Resource Allocation

From a health policy perspective, recognizing the differential impact of planned and unplanned hospitalizations has implications for resource allocation. Investing in planned hospitalizations and structured programs may prove cost-effective by reducing the need for resource-intensive unplanned hospitalizations. Reimbursement models should consider supporting planned hospitalizations for HF education and rehabilitation as part of standard HF management to optimize long-term cost savings and patient outcomes.

### Prior Evidence on Hospitalization Burden

Our results align with large registry and economic analyses showing that hospitalizations dominate direct HF costs and that unplanned hospitalizations are particularly expensive and prognostically unfavorable.[2, 4, 37] The present analysis extends prior work by directly comparing planned versus unplanned hospitalizations within the same patients, demonstrating materially lower per-hospitalization costs and shorter stays for planned hospitalizations.

### Limitations and Future Directions

First, this was a single-center, retrospective study with a small cohort (n=20), which limits generalizability and precludes robust risk-adjusted modeling. Further, differences in reimbursement systems across countries should also be taken into account. Also, selection bias and residual confounding (e.g., socioeconomic factors, outpatient adherence, community resources) cannot be excluded. Second, the content and structure of the Kurume-HEARTS program are not yet standardized and are currently being implemented in an exploratory manner. Third, we did not assess long-term outcomes such as all-cause mortality, cardiovascular mortality, or health-related QoL. To address these limitations, we have initiated prospective, multicenter studies.

## Conclusions

The Kurume-HEARTS program can help reduce the cost and frequency/length of unplanned admissions by enabling earlier intervention and structured inpatient management (**Graphical abstract**). Proactive use of planned hospitalizations—focused on structured education, cardiac rehabilitation, and GDMT optimization—may mitigate the clinical and economic burden of recurrent decompensation.

In addition, in Japan, there is a growing need to foster and expand the roles of registered instructor of cardiac rehabilitation (The Japanese Association of Cardiac Rehabilitation) and certified HF educators (The Japanese Circulation Society). The Kurume-HEARTS program may serve as an effective platform for such professionals to apply and further develop their expertise, potentially creating a synergistic effect that enhances the overall quality of HF care.

## Acknowledgments

We thank all medical staffs who work in the Department of Cardiovascular Medicine and the Cardiac Care Unit at Kurume University Hospital.

## Data availability statement

The data underlying this article will be shared with others for the purpose of reproducing results or replicating procedures upon reasonable request to the corresponding author, participant to institutional and ethics committee approval.

## Authors’ Contributions

Toshiyuki Yanai: Conceptualization, Methodology, Formal analysis, Data Curation, and Writing-Original draft preparation.

Tatsuhiro Shibata: Methodology, Formal analysis, Investigation, Writing-Review and editing.

Kodai Shibao: Methodology, Investigation.

Daiki Akagaki: Methodology, Investigation.

Kota Okabe: Methodology, Investigation.

Shoichiro_Nohara: Methodology, Investigation, Writing-Review and editing.

Jinya Takahashi: Methodology, Investigation, Writing-Review and editing.

Koutatsu Shimozono: Methodology, Data Curation, Writing-Review, and Formal analysis.

Yoshihiro Fukumoto: Project administration, Conceptualization, Writing-Review and editing.

## Abbreviations list

ADHF: acute decompensated heart failure
ANOVA: analysis of variance
BMI: body mass index
CRT-D: cardiac resynchronization therapy defibrillator
eGFR: estimated glomerular filtration rate
GDMT: guideline-directed medical therapy
GMR: geometric mean ratio
HF: heart failure
JCS: Japanese Circulation Society
JHFS: Japanese Heart Failure Society
LVEF: left ventricular ejection fraction
NT-proBNP: N-terminal pro-B-type natriuretic peptide
NYHA: New York Heart Association
LDL-C: low-density lipoprotein cholesterol
MRA: mineralocorticoid receptor antagonist
QoL: quality of life
RAAS: renin-angiotensin-aldosterone system
SD: standard deviation
SGLT2: sodium-glucose cotransporter 2.

